# CSF inflammatory cytokines as prognostic indicators for cognitive decline across Alzheimer’s disease spectrum

**DOI:** 10.1101/2024.11.13.24317270

**Authors:** Elham Ghanbarian, Babak Khorsand, Kellen K. Petersen, Bhargav T. Nallapu, S. Ahmad Sajjadi, Richard B. Lipton, Ali Ezzati

## Abstract

**Background:** A growing body of evidence suggests that neuroinflammation contributes actively to pathophysiology of Alzheimer’s disease (AD) and promotes AD progression. The predictive value of neuroinflammatory biomarkers for disease-staging or estimating disease progression is not well understood. In this study, we investigate the diagnostic and prognostic utility of inflammatory biomarkers in combination with conventional AD biomarkers.

**Methods:** We included 258 participants from the Alzheimer’s Disease Neuroimaging Initiative (ADNI) who had CSF biomarkers of β-Amyloid (Aβ), tau, and inflammation. The primary outcome of interest was clinically meaningful cognitive decline (CMCD) as defined by an increase of ≥4 on the Alzheimer’s Disease Assessment Scale Cognitive Subscore 11 (ADAS-11, scores 0-70, higher scores indicate worse cognition). Predictor variables were categorized as demographics (D; age, sex, and education), genetic (APOE4 status (A)), inflammatory biomarkers (I), and classic (C) cerebrospinal fluid (CSF) biomarkers of Aβ and p-tau181. Simultaneous inclusion of eleven CSF inflammatory biomarkers as covariates in logistic regression models was examined to assess improvements in classifying baseline clinical diagnoses (cognitively normal (CN), mild cognitive impairment (MCI), Dementia) and predicting individuals with and without CMCD over one year of follow-up.

**Results:** At 1-year follow up, 27.1% of participants experienced CMCD. Inclusion of inflammatory biomarkers improved baseline classification of CN vs MCI as well as CN vs Dementia for models including D and A variables (DA; both *p*<0.001). Similarly, when classic CSF biomarkers of AD were included into the model (DAC model), inclusion of inflammatory markers improved classification of CN vs MCI (*p*<0.01) as well as CN vs Dementia (*p*<0.001). Addition of inflammatory biomarkers to both DA and DAC models improved predictive performance for CMCD in persons with baseline MCI and Dementia (all *p*<0.05), but not in the CN group. In addition, the predictive performance of the DAI model was superior to the DAC model in the MCI and Dementia groups (both *p*<0.05).

**Conclusions:** Addition of CSF inflammatory biomarkers to CSF biomarkers of AD can improve diagnostic accuracy of clinical disease stage at baseline and add incremental value to AD biomarkers for prediction of cognitive decline.

**Learning objective:** □ Identify potential diagnostic and prognostic ability of inflammatory biomarkers to assess Alzheimer’s disease stage and cognitive decline.

## 1 Introduction

Alzheimer’s disease (AD) is widely acknowledged as a complex condition, characterized by heterogeneous underlying pathologies, which can manifest differently across individuals. Neuroinflammation is known to play an important role in the pathogenesis of AD.^1^ It actively contributes to formation of the main AD pathological hallmarks of amyloid plaques and neurofibrillary tangles.^2^ Early in the course of AD pathology, brain immune cells, such as microglia and astrocytes, become activated^3^ and are involved in the production and clearance of β-Amyloid (Aβ) and tau, and their interaction with amyloid plaques accelerates neurodegeneration.^4, 5^

Biomarkers of neuroinflammation have been shown to be elevated in the cerebrospinal fluid (CSF) at various stages of the AD continuum, including the preclinical (cognitively normal, CN), prodromal (mild cognitive impairment, MCI), and dementia stages.^3, 6, 7^ Therefore, incorporating these biomarkers into clinical assessments has the potential to enhance the accuracy of diagnosis and disease staging. One study has demonstrated that the inclusion of CSF inflammatory biomarkers in classifiers improves the differentiation of AD from other forms of dementia as well as non-demented groups^8^.

However, the number of studies supporting this approach remains very limited. Moreover, recent studies suggest that neuroinflammatory responses contribute to the acceleration of AD progression. Current evidence indicates that higher baseline levels of CSF neuroinflammatory markers are associated with an increased risk of progression to MCI and AD, and including these biomarkers in predictive models significantly improves the accuracy of predicting incident dementia. ^6, 9, 10, 11^

The diagnostic and prognostic value of the core biomarkers of AD in determining disease progression across the AD spectrum is previously studied by our group,^12,13^ as well as others.^14, 15^ However, studies examining the diagnostic and predictive value of neuroinflammatory markers are limited. In this study, we used data from the Alzheimer’s Disease Neuroimaging Initiative (ADNI) to investigate whether integrating baseline neuroinflammatory biomarkers with AD-specific biomarkers of Aβ and phosphorylated tau (p-tau) improves the ability for classification of clinical stage (i.e., diagnostic accuracy) and the ability to predict disease progression (prognostic performance) over a one-year follow-up period.

## 2 Methods

### 2.1 Participants

#### 2.1.1. ADNI study design

The data used in the preparation of this article were obtained from the ADNI database (adni.loni.usc.edu). The ADNI was launched in 2003 as a public-private partnership, led by Principal Investigator Michael W. Weiner, MD, with the primary goal of testing whether magnetic resonance imaging (MRI), positron emission tomography (PET), biological markers, and clinical and neuropsychological assessment can be combined to measure the progression of mild cognitive impairment (MCI) and early AD. ADNI participants were selected from a convenience sample, and at enrollment, they were aged between 55–90 (inclusive), classified into cognitively normal (CN), MCI, or dementia diagnosed individuals. They underwent periodic evaluations of functional, biomedical, neuropsychological, and clinical assessments. ADNI data collection was approved by the institutional review boards of all participating institutions. Informed written consent was obtained from all participants at each site. Detailed information on measures and methods of assessment in the ADNI project are available at http://www.adni.loni.usc.edu.

#### 2.1.2. Current study participants

For this study, participants were selected who meet specific inclusion criteria: They had to have CSF biomarkers of Aβ, phosphorylated tau 181 (p-tau181), and inflammation markers at baseline visit. Additionally, participants needed to have a minimum of 1 year of follow-up data on cognitive performance.

#### 2.1.3. Diagnostic classification

CN individuals had Mini-Mental-State Examination (MMSE) scores of 24 or higher; Clinical Dementia Rating (CDR) score of 0 with no evidence of depression and no memory complaints. MCI was defined as MMSE score between 24 and 30, inclusive; CDR score of 0.5 with report of memory complaints and no significant functional impairment. All individuals with MCI also met Petersen criteria. Participants with dementia met National Institute of Neurological and Communicative Disorders and Stroke–Alzheimer’s Disease and Related Disorders Association (NINCDS-ADRDA) criteria for clinically defined probable AD, and have MMSE scores between 20 and 26 (inclusive), and CDR of 0.5 or 1.^16^

### 2.2. Study measures and outcomes

#### 2.2.1 CSF biomarkers

##### AD biomarkers

CSF biomarkers of Aβ42 and p-tau181 measured at the ADNI Biomarker Core Laboratory (University of Pennsylvania) using the multiplex xMAP Luminex platform and Innogenetics/Fujirebio AlzBio3 immunoassay reagents, details of which have been previously described.^17^

##### Inflammatory biomarkers

Blinded CSF samples that were transported from Biomarker Core to Emory University were examined by William Hu and J. Christina Howell in the Department of Neurology. Commercially available multiplex immunoassays (Millipore Sigma, Burlington, MA, USA) were used to measure all biomarkers in duplicate. Details for measurement of these biomarkers are provided elsewhere^14^. Eleven CSF inflammatory biomarkers were considered for this study: Tumor necrosis factor receptor 1 (TNFR1), TNFR2, Transforming growth factor beta 1 (TGF-β1), TGF-β2, TGF-β3, Interleukin-6 (IL-6), IL-7, IL-10, IL-21, Tumor Necrosis Factor alpha (TNF-α), and Intercellular Adhesion Molecule 1 (ICAM-1). Inflammatory biomarkers were classified into three larger groups: anti-inflammatory (TGF-β1), TGF-β2, TGF-β3, IL-10), pro-inflammatory (TNFR1, TNF-α, IL-6, IL-21, ICAM-1) and mixed-inflammatory (TNFR2, IL-7) biomarkers. ^1, 18^

#### 2.2.3. APOE gene status

Apolipoprotein E (APOE) ε4 allele frequency was available for all participants included in this study. ApoE4 status was defined as ApoE4 negative (–) if they carried no ApoE4 allele or ApoE4 positive (+) if they carried at least one ApoE4 allele.

#### 2.2.2. Cognitive outcomes

The primary neuropsychological test used for this study was Alzheimer’s Disease Assessment Scale cognitive subscale 11 (ADAS-11).^19^ Originally designed as an outcome measure for dementia intervention trials, the ADAS-11 serves as a primary indicator of global cognition in assessing the response to antidementia therapies. ADAS-11 raw scores range from 0 to 70, where higher scores indicate greater cognitive dysfunction. Our primary outcome of interest was clinically meaningful cognitive decline (CMCD), defined by an increase of ≥4 on the ADAS-11 one year after the baseline assessment.^20^

### 2.3 Statistical analysis

Differences in participant characteristics between individuals with stable cognition and meaningful cognitive decline were tested using t-test for continuous variables and chi square test for categorical variables. Logistic regression models were employed to develop classification and predictive models. These models addressed two primary questions: first, whether simultaneous inclusion of eleven CSF inflammatory biomarkers, alongside demographics, APOE4 genotype and CSF biomarkers of AD, improves the accuracy of diagnostic classification. The models were used to classify individuals into three paired groups: CN vs. MCI, CN vs. Dementia, and MCI vs. Dementia. The second question assessed whether the addition of all eleven inflammatory biomarkers to other variables would improve the prediction of CMCD within a one-year timeframe. Therefore, the outcome of interest was defined as one of two following groups based on data from 1-year follow up: individuals with CMCD versus those without CMCD. The base model (DA) included demographics including age, sex, and education (D), and the genetic risk factor of APOE4 (A). For the second model (DAC), classic CSF AD biomarkers of Aβ and p-tau181 (C) were added to the base model. Finally, eleven CSF inflammatory biomarkers (I) were simultaneously added to both models, resulting in DAI and DACI models.

DeLong’s test was used to compare the Area Under the Curves (AUCs) from logistic regression models for binary classification of baseline clinical diagnosis with and without the inflammatory biomarkers, and for predicting clinically meaningful cognitive decline at 1 year. To determine the importance of different variables in classifying the outcome, we followed methods previously used for feature ranking^21^ with some modifications. Each variable was excluded from the logistic regression models, and the resulting decrease in AUC was compared to the AUC with all features included. The greater the decrease in AUC, the more important the variable was in the model.

## 3 Results

### 3.1 Baseline Characteristics

Table 1 provides a summary of the baseline characteristics of the entire study population, as well as for the two subgroups classified by change in cognitive performance after one year. A total of 258 participants were included in this study: 74 (28.7%) CN, 122 (47.3%) MCI and 62 (24.0%) with Dementia diagnosis at baseline. Participants had 15.78 (SD 2.98) years of education, 41.9% were women, and 52.7% were *APOE ε4* carriers. After one year from baseline, 70 participants (27.1%) had CMCD (progressors), and 188 participants (72.9%) remained cognitively stable. The progressor subgroup compared to the stable subgroup included more females (57.1% vs 36.2%, *p*<0.01), had lower levels of CSF Aβ pathology (*p*<0.01), and higher p-tau181 (*p*<0.001). CSF levels of inflammatory biomarkers were not significantly different between the two subgroups.

**Table 1.**
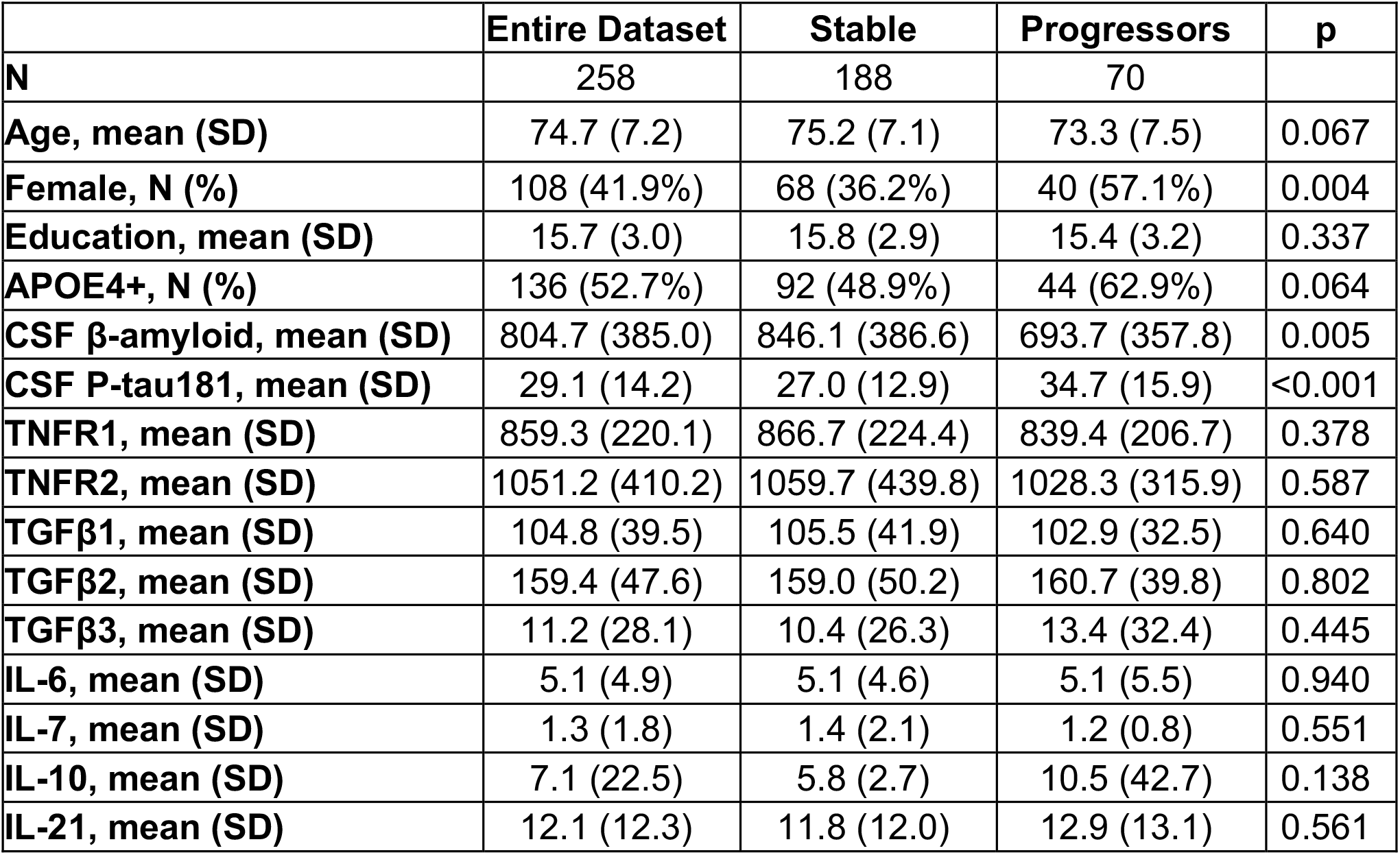
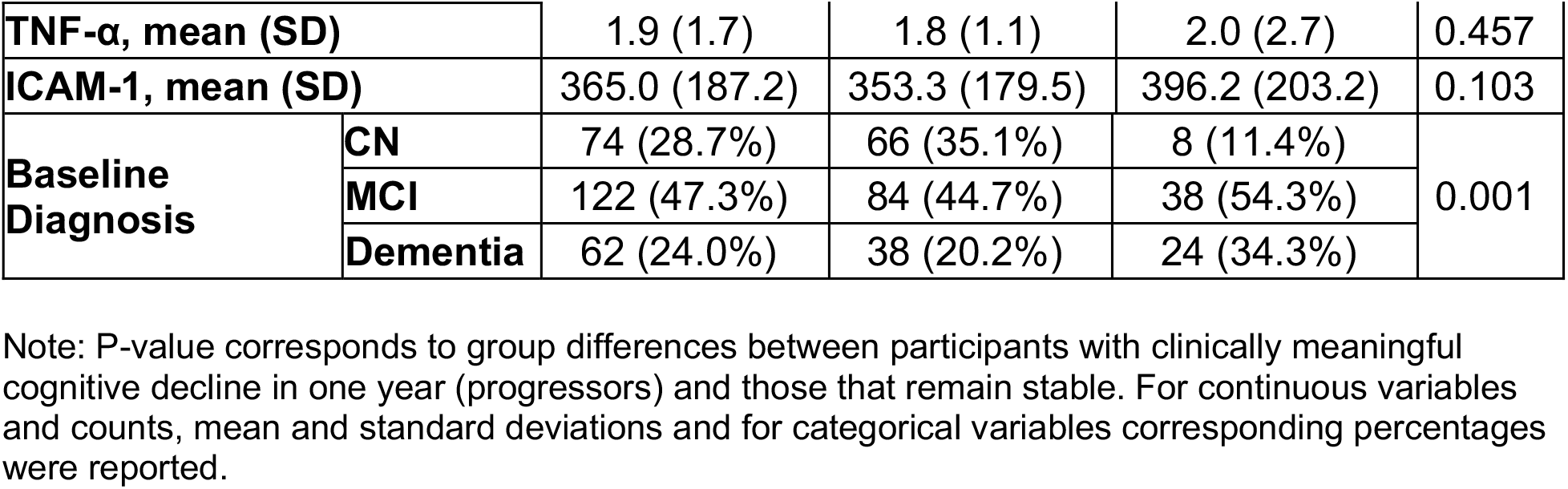
Characteristics of the entire sample and stratified by stable cognition and cognitive decline (progressors) over one year in terms of clinically meaningful change (≥ 4 points) in ADAS-11.

|  | Entire Dataset | Stable | Progressors | p |
| --- | --- | --- | --- | --- |
| **N** | 258 | 188 | 70 |  |
| **Age, mean (SD)** | 74.7 (7.2) | 75.2 (7.1) | 73.3 (7.5) | 0.067 |
| **Female, N (%)** | 108 (41.9%) | 68 (36.2%) | 40 (57.1%) | 0.004 |
| **Education, mean (SD)** | 15.7 (3.0) | 15.8 (2.9) | 15.4 (3.2) | 0.337 |
| **APOE4+, N (%)** | 136 (52.7%) | 92 (48.9%) | 44 (62.9%) | 0.064 |
| **CSF β-amyloid, mean (SD)** | 804.7 (385.0) | 846.1 (386.6) | 693.7 (357.8) | 0.005 |
| **CSF P-tau181, mean (SD)** | 29.1 (14.2) | 27.0 (12.9) | 34.7 (15.9) | <0.001 |
| **TNFR1, mean (SD)** | 859.3 (220.1) | 866.7 (224.4) | 839.4 (206.7) | 0.378 |
| **TNFR2, mean (SD)** | 1051.2 (410.2) | 1059.7 (439.8) | 1028.3 (315.9) | 0.587 |
| **TGFβ1, mean (SD)** | 104.8 (39.5) | 105.5 (41.9) | 102.9 (32.5) | 0.640 |
| **TGFβ2, mean (SD)** | 159.4 (47.6) | 159.0 (50.2) | 160.7 (39.8) | 0.802 |
| **TGFβ3, mean (SD)** | 11.2 (28.1) | 10.4 (26.3) | 13.4 (32.4) | 0.445 |
| **IL-6, mean (SD)** | 5.1 (4.9) | 5.1 (4.6) | 5.1 (5.5) | 0.940 |
| **IL-7, mean (SD)** | 1.3 (1.8) | 1.4 (2.1) | 1.2 (0.8) | 0.551 |
| **IL-10, mean (SD)** | 7.1 (22.5) | 5.8 (2.7) | 10.5 (42.7) | 0.138 |
| **IL-21, mean (SD)** | 12.1 (12.3) | 11.8 (12.0) | 12.9 (13.1) | 0.561 |

|  |  |  |  |  |  |
| --- | --- | --- | --- | --- | --- |
| **TNF-α, mean (SD)** |  | 1.9 (1.7) | 1.8 (1.1) | 2.0 (2.7) | 0.457 |
| **ICAM-1, mean (SD)** |  | 365.0 (187.2) | 353.3 (179.5) | 396.2 (203.2) | 0.103 |
| **Baseline Diagnosis** | **CN** | 74 (28.7%) | 66 (35.1%) | 8 (11.4%) | 0.001 |
|  | **MCI** | 122 (47.3%) | 84 (44.7%) | 38 (54.3%) |  |
|  | **Dementia** | 62 (24.0%) | 38 (20.2%) | 24 (34.3%) |  |

Note: P-value corresponds to group differences between participants with clinically meaningful cognitive decline in one year (progressors) and those that remain stable. For continuous variables and counts, mean and standard deviations and for categorical variables corresponding percentages were reported.

### 3.2. Clinical disease stage classification

Receiver Operating Characteristic (ROC) curves of logistic regression models used in binary classification of baseline clinical diagnosis are shown in Figure 1, and the corresponding Area Under the Curves (AUCs) in Table 2. Simultaneous inclusion of eleven inflammatory biomarkers in the DA model improved the classification of clinical stage diagnosis between CN versus MCI (AUC 0.81 vs 0.70, *p < 0*.*01)* and CN versus Dementia (AUC 0.91 vs 0.79, *p < 0*.*001)* but not between MCI versus Dementia. The same results were obtained when all inflammatory biomarkers were added to the DAC model; they improved the classification of diagnosis between CN versus MCI (AUC 0.84 vs 0.74, *p < 0*.*01*) and CN versus Dementia (AUC 0.94 vs 0.83, *p < 0*.*001*) but not between MCI versus Dementia.

**Table 2.** Area Under the Curve (AUC) from logistic regression models in binary classification of baseline clinical diagnosis, comparing models with and without inflammatory biomarkers (I). Basic models include demographics (D: age, sex, and education), APOE4 status (A), and Classic AD biomarkers (C: CSF Aβ and p-tau181). P-values correspond to DeLong’s test applied to the classifiers. Eleven CSF inflammatory biomarkers were considered as a group (all or none were included): TNFR1, TNFR2, TGFβ1, TGFβ2, TGFβ3, IL-21, IL-6, IL-7, IL-10, TNF-α, and ICAM-1.

| Binary Classification | Inflammatory Biomarkers (I) | DA Model |  | DAC Model |  |
| --- | --- | --- | --- | --- | --- |
|  |  | AUC (95% CI) | p | AUC (95% CI) | p |
| CN vs MCI | without I | 0.70 (0.63 - 0.78) | 0.001 | 0.74 (0.63 - 0.78) | 0.002 |
|  | with I | 0.81 (0.74 - 0.87) |  | 0.84 (0.78 - 0.89) |  |
| MCI vs Dementia | without I | 0.66 (0.57 - 0.74) | 0.35 | 0.71 (0.60 - 0.76) | 0.33 |
|  | with I | 0.68 (0.63 - 0.79) |  | 0.73 (0.66 - 0.80) |  |
| CN vs Dementia | without I | 0.79 (0.71 - 0.87) | 0.0004 | 0.83 (0.86 - 0.96) | 0.0001 |
|  | with I | 0.91 (0.77 - 0.90) |  | 0.94 (0.91 - 0.98) |  |

**Figure 1.**
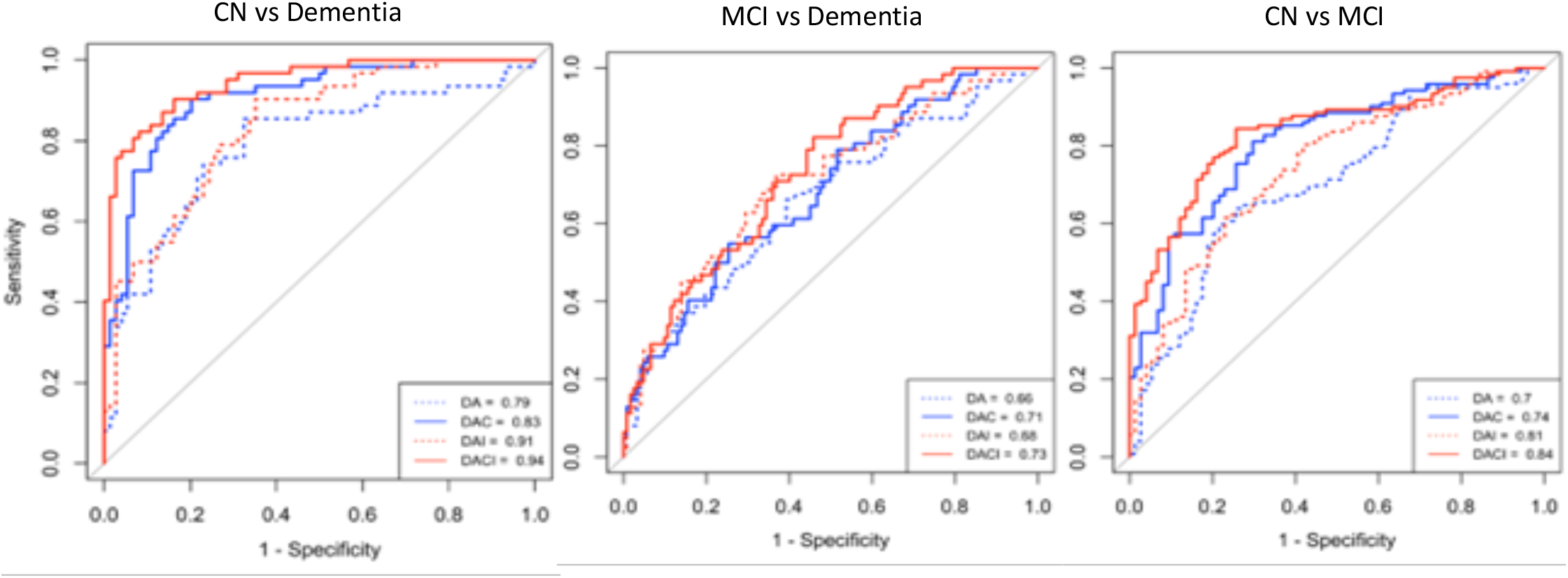
Receiver operating characteristic (ROC) curves from logistic regression in binary classification of baseline clinical diagnosis comparing models with and without inflammatory biomarkers (I). Basic models include demographics (D: age, sex, and education), APOE4 status (A), and Classic AD biomarkers (C: CSF Aβ and p-tau181). Blue colors represent ROC curves for models without inflammatory biomarkers. Red colors represent ROC curves with inflammatory biomarkers. The legend includes Area Under the ROC Curve (AUC) for each model.

### 3.3. Prediction of CMCD

ROC curves of logistic regression models used in predicting clinically meaningful cognitive decline at 1 year and corresponding AUCs in the forest plot are shown in Figure 2. Inclusion of inflammatory biomarkers in the DA model (DA vs DAI) significantly improved its predictive performance for both the MCI (AUC 0.8 vs 0.65, *p < 0*.*05*) and Dementia (AUC 0.81 vs 0.62, *p < 0*.*05*) groups, but not in the CN group. Similarly, the inclusion of inflammatory biomarkers in the DAC model (DAC vs DACI) significantly improved the model’s predictive performance in the MCI (AUC 0.81 vs 0.68, *p < 0*.*05*) and Dementia (AUC 0.81 vs 0.64, *p < 0*.*05*) groups, but not in the CN group. In addition, the predictive performance of the DAI model was superior to the DAC model in both the MCI (AUC 0.8 vs 0.68, *p* < 0.05) and Dementia (0.81 vs 0.64, *p* < 0.05) groups.

**Figure 2.**
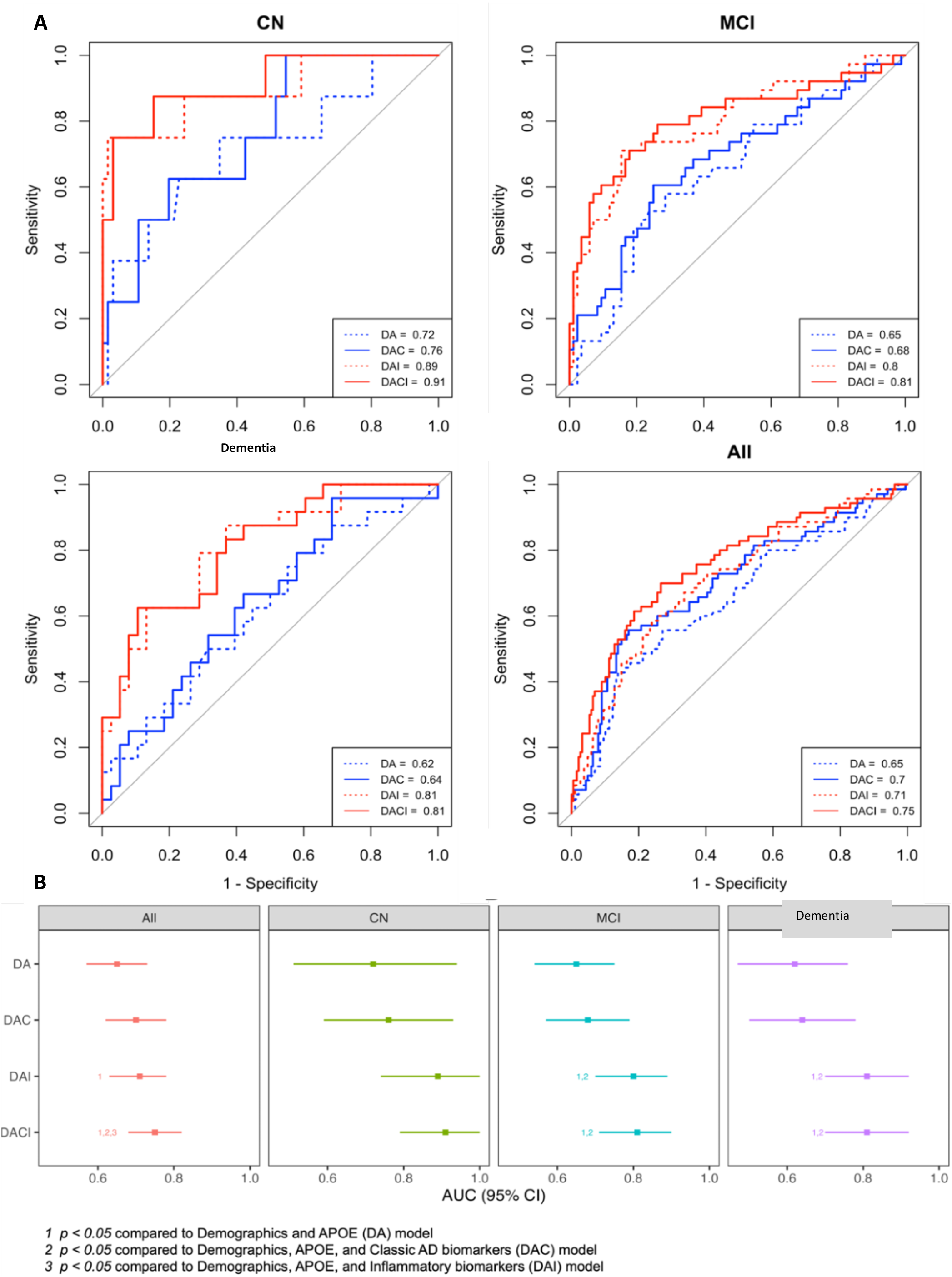
**A**. Receiver operating characteristic (ROC) curves from logistic regression for predicting meaningful cognitive decline (≥4 points change in ADAS-11) at 1 year, for all participants and stratified by baseline diagnosis, for models with and without inflammatory biomarkers (I). Basic models include demographics (D: age, sex, and education), APOE4 status (A), and Classic AD biomarkers (C: CSF Aβ and p-tau181). Blue and red colors represent ROC curves for models without and with inflammatory biomarkers, respectively. The legend includes Area Under the ROC Curve (AUC) for each model. **B**. Forest plot for the Area Under the ROC Curve (AUC) from logistic regression for predicting meaningful cognitive decline (≥4 points change in ADAS-11) at 1 year, for all participants and stratified by baseline diagnosis for models with and without inflammatory biomarkers (I). Basic models include demographics (D: age, sex, and education), APOE4 status (A), and Classic AD biomarkers (C: CSF Aβ and p-tau181). DeLong’s tests were used to compare the AUCs, and the footnotes below the figure show which comparisons were significantly different with *P* < 0.05.

### 3.4. Feature importance in predictive models

To identify feature sets with the highest predictive value for meaningful cognitive decline at 1-year, inflammatory biomarkers were categorized into three groups: anti-inflammatory (TGF-β1, TGF-β2, TGF-β3 & IL-10), pro-inflammatory (TNFR1, TNF-α, IL-6, IL-21 & ICAM-1), and mixed-inflammatory (TNFR2 & IL-7) biomarkers. Then, using the DACI model, we excluded each group from the model and measured the resulting decrease in the AUC (Figure 3). In the CN and MCI groups, excluding pro-inflammatory biomarkers resulted in the largest decrease in the AUC (9.1% and 8.7%, respectively), while in the Dementia group, anti-inflammatory biomarkers had the largest impact (5.2%).

**Figure 3.**
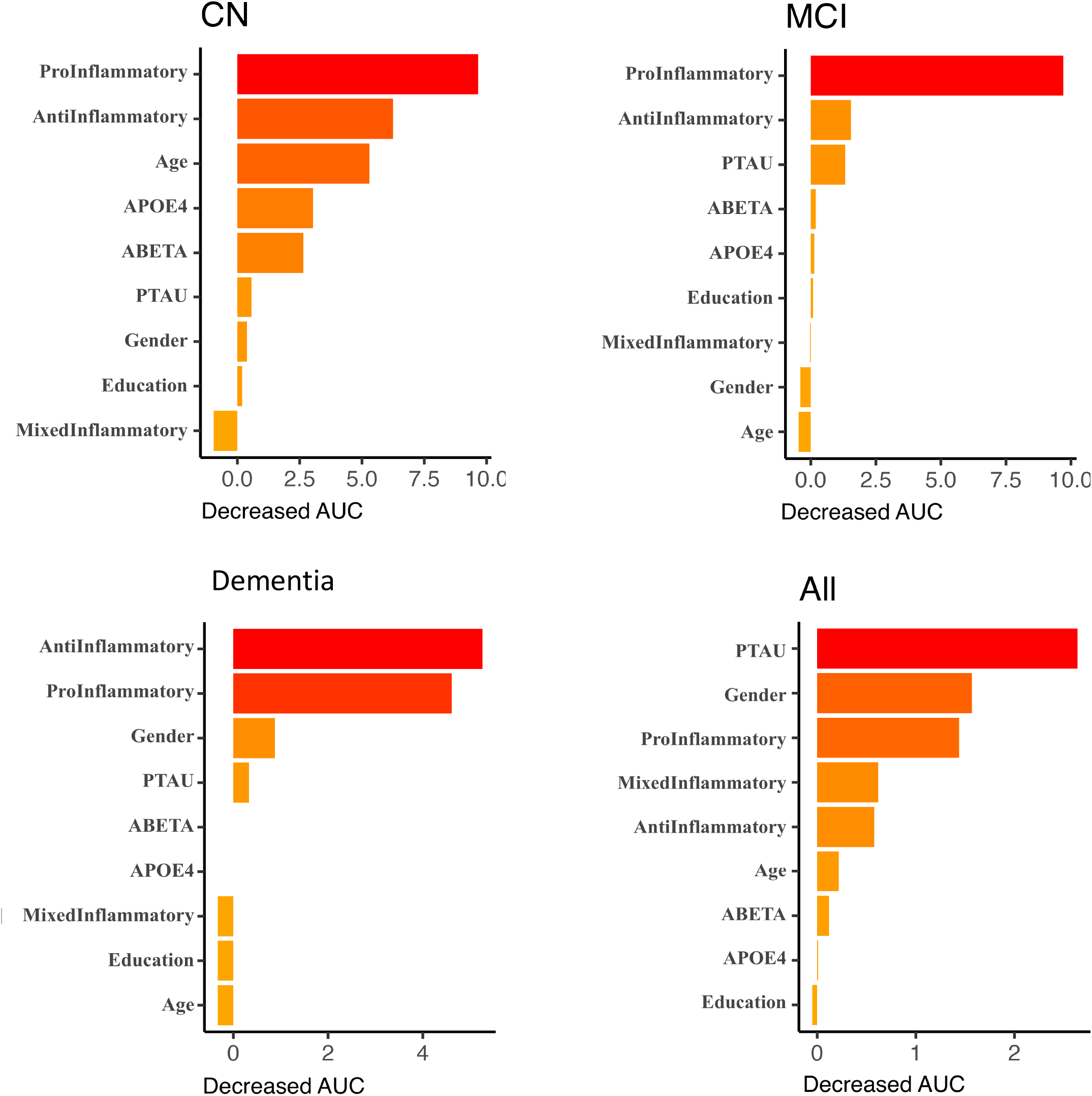
Feature importance plot for predicting meaningful cognitive decline (≥4 points change in ADAS-11) at 1 year, for all participants and stratified by baseline diagnosis. CSF inflammatory biomarkers are classified into 3 categories: Pro-inflammatory (TNFR1, TNF-α, IL-6, IL-21, and ICAM-1), Anti-inflammatory (TGFβ1, TGFβ2, TGFβ3, and IL-10), and Mixed inflammatory (TNFR2 and IL-7).

## 4 Discussion

We found that inclusion of CSF inflammatory biomarkers, along with classic biomarkers of AD, significantly improved our ability for clinical disease staging, particularly for differentiating between CN individuals and those with MCI, as well as between CN and dementia groups. Furthermore, the predictive performance of models for disease progression in individuals with MCI and dementia showed improvement over a one-year period. Notably, pro-inflammatory markers were more important in predicting short term cognitive decline in the CN and MCI groups, while anti-inflammatory markers played a more significant role in the dementia group.

Inflammatory biomarkers are not currently used for diagnosing or staging of AD^22^; however, emerging evidence suggest that inclusion of these biomarkers can enhance both clinical diagnosis and disease staging accuracy. It has been shown that inclusion of CSF inflammatory biomarkers, including IL-7 and IL-10, improves the diagnosis of AD versus frontotemporal dementia and non-demented groups in a small population study with 68 subjects.^8^ Evidence shows that CSF neuroinflammatory markers correlate with baseline AD pathology and cognitive function across the AD continuum. It has been reported that CSF levels of several cytokines, including ICAM-1, were already elevated at the preclinical (CN) and prodromal (MCI) stages of AD, with higher levels of these biomarkers being associated with increased levels of CSF p-tau and t-tau as well as worse cognitive performance.^6, 23^ In patients with MCI, higher IL-6 levels have been associated with increased baseline white matter lesion, suggesting that inflammation may contribute to early neural damage and cognitive decline in the prodromal stage.^24^ Similarly, in patients already diagnosed with AD, several CSF inflammatory markers have been negatively correlated with baseline cognitive function, further supporting the role of neuroinflammation in disease progression at this stage.^8, 25^ Therefore, inclusion of these biomarkers can potentially improve baseline diagnosis and staging of AD, as shown in our findings.

It is widely accepted that inflammation plays a significant role in the pathophysiology of AD, even in the early asymptomatic stages. However, studies that have evaluated the relationship between CSF inflammatory markers and incidence of cognitive decline over time have yielded mixed and often contradictory findings. Several studies indicate that elevated levels of neuroinflammatory markers are associated with higher risk of progression to MCI or dementia. In a large longitudinal study with 821 subjects, baseline CSF inflammatory markers, including ICAM-1, were associated with a more rapid cognitive decline over time.^6^ Notably, these biomarkers were independent predictors of time to AD diagnosis, with the highest levels of ICAM-1 linked to a significantly increased risk of AD compared to the lowest levels. Another study demonstrated that elevated baseline CSF cytokines, such as IL-9 and TNF-α, were associated with a higher risk of progression to MCI and AD, particularly in females and APOE carriers.^9^ Also, in MCI patients who subsequently developed AD, CSF (and plasma) levels of baseline TNF-α receptors (TNFR1 and TNFR2) were higher compared to MCI subjects who remained cognitively stable as well as aged-matched healthy controls.^10^ Interestingly, the annual cognitive decline during 4-6 years of follow-up was correlated to the baseline levels of these markers both in CSF and plasma. A key finding in our study is that incorporating CSF inflammatory markers significantly enhanced the predictive performance of the models (DAI vs. DA and DACI vs. DAC). Notably, when comparing the DAC and DAI models, the latter demonstrated superior overall performance, suggesting that, in the short term (i.e., 1 year), the predictive value of neuroinflammatory biomarkers may surpass that of traditional AD biomarkers. This observation warrants further investigation in other cohorts, ideally with more frequent biomarker assessments and cognitive outcome measurements within the same timeframe.

On the other hand, several studies have shown an inverse relationship between CSF inflammatory markers and cognitive decline. A large population study (including 742 individuals) with 8 years of follow-up showed that higher baseline levels of CSF cytokines, such as IL-8 and IL-16, were associated with more extensive white matter lesions at baseline in both CN and MCI individuals.^24^ However, unexpectedly, lower levels of these cytokines were associated with increased white matter lesions over time in MCI patients. More importantly, regarding cognitive outcome, in the CN group, higher baseline CSF cytokine levels (such as IL-8 and sICAM-1) were associated with greater longitudinal cognitive decline, whereas no significant associations were found in the MCI group. Similarly, in a small study with 32 AD patients, several CSF cytokines including IL-6 and IL-10 have shown negative association with cognitive decline after 1 year.^8^ In fact, these findings indicate that a stronger inflammatory response might result in better clinical outcomes, possibly because inflammation is associated with the clearance of toxic proteins.^1^

Similarly, systemic markers of inflammation have demonstrated significant yet inconsistent effects in different studies. In the preclinical stage of AD, higher plasma IL-6 levels have been shown to predict subsequent conversion to MCI, as well as increased global and regional Aβ deposition over a 2-years follow up period.^26^ In contrast, in a study on ADNI dataset, lower levels of plasma cytokines such as IL-6 and IL-16 have been reported in patients with MCI compared to CN group, and higher plasma IL-6 receptor levels significantly decreased the risk of progression from MCI to dementia over 2 years of follow-up.^27^ Also, it has been reported that increase in some plasma cytokine levels (IL-18) are greater in patients with mild AD than in those with advanced AD.^28, 29^ The conflicting results across studies can be attributed to differences in cohort characteristics, the selection of CSF versus plasma biomarkers, inclusion of single versus grouped inflammatory markers in the predictive models, the impact of other concurrent systemic and inflammatory diseases, the inclusion of pro-inflammatory versus anti-inflammatory markers, the stage of the disease within the AD continuum, and also, the age of the patients.^30^

We also found that pro-inflammatory markers were more important in predicting short term cognitive decline in the CN and MCI groups, while anti-inflammatory markers were more critical in the dementia group. This shift from pro-to anti-inflammatory marker may reflect a dynamic inflammatory response as AD progresses. This is consistent with recent evidence indicating that differences in inflammatory expression profiles may vary based on the disease state.^28, 31^ Early in the disease, pro-inflammatory cytokines released by microglia might dominate as immune system counteracts accumulation of amyloid plaques and tau tangles. However, in the more advanced stages, an anti-inflammatory response could indicate a failing compensatory mechanism as the disease reaches a chronic neuroinflammation and neuronal damage.^1, 32^

A few limitations should be acknowledged. We lacked access to a comparable dataset from a secondary source to validate our findings. Enhancing access to data collected in research studies will enable us to obtain additional datasets, allowing us to further improve this line of work and develop well-validated models. All inflammatory biomarkers were measured at a single time point, potentially impacted by other ongoing systemic or central inflammatory processes. It is well established that systemic diseases, such as infections or rheumatologic disorders, can influence the levels of inflammatory biomarkers both centrally and peripherally. However, due to the lack of data on these potential confounders, we were unable to control for them in our analysis. Despite these limitations, it is encouraging that we found a robust impact of inflammatory markers for diagnostic and prognostic purposes.

In conclusion, our study demonstrates that the inclusion of CSF inflammatory biomarkers, alongside established AD biomarkers, improves both diagnostic accuracy and predictive power for cognitive outcomes in MCI and dementia. These findings highlight the potential of neuroinflammatory markers as a valuable component in the clinical assessment and management of AD.

## Data Availability

https://adni.loni.usc.edu/
we used ADNI data which is openly available here.

## Acknowledgment

This study was supported in part by grants from the National Institute of Health (NIA K23 AG063993; AG080635; AG003949), the Alzheimer’s Association (SG-24-988292 ISAVRAD), Cure Alzheimer’s Fund, and “the Leonard and Sylvia Marx Foundation”.

